# Identification of social engagement indicators associated with autism spectrum disorder using a game-based mobile application

**DOI:** 10.1101/2021.06.20.21259187

**Authors:** Maya Varma, Peter Washington, Brianna Chrisman, Aaron Kline, Emilie Leblanc, Kelley Paskov, Nate Stockham, Jae-Yoon Jung, Min Woo Sun, Dennis P. Wall

**Affiliations:** Departments of Computer Science, Stanford University, Stanford, CA, USA; Departments of Biomedical Data Science, Stanford University, Stanford, CA, USA; Departments of Neuroscience, Stanford University, Stanford, CA, USA; Departments of Bioengineering, Stanford University, Stanford, CA, USA; Departments of Pediatrics, Stanford University, Stanford, CA, USA

**Keywords:** mobile health, autism spectrum disorder, social phenotyping, computer vision

## Abstract

**Objective:** Autism spectrum disorder (ASD) is a widespread neurodevelopmental condition with a range of potential causes and symptoms. Children with ASD exhibit behavioral and social impairments, giving rise to the possibility of utilizing computational techniques to evaluate a child’s social phenotype from home videos.

**Methods:** Here, we use a mobile health application to collect over 11 hours of video footage depicting 95 children engaged in gameplay in a natural home environment. We utilize automated dataset annotations to analyze two social indicators that have previously been shown to differ between children with ASD and their neurotypical (NT) peers: (1) gaze fixation patterns and (2) visual scanning methods. We compare the gaze fixation and visual scanning methods utilized by children during a 90-second gameplay video in order to identify statistically-significant differences between the two cohorts; we then train an LSTM neural network in order to determine if gaze indicators could be predictive of ASD.

**Results:** Our work identifies one statistically significant region of fixation and one significant gaze transition pattern that differ between our two cohorts during gameplay. In addition, our deep learning model demonstrates mild predictive power in identifying ASD based on coarse annotations of gaze fixations.

**Discussion:** Ultimately, our results demonstrate the utility of game-based mobile health platforms in quantifying visual patterns and providing insights into ASD. We also show the importance of automated labeling techniques in generating large-scale datasets while simultaneously preserving the privacy of participants. Our approaches can generalize to other healthcare needs.

## 1. Introduction

Autism spectrum disorder (ASD) is a neurodevelopmental disorder characterized by social impairments, communication difficulties, and restricted and repetitive patterns of behavior. Currently, 1 in 59 children in the United States have been diagnosed with ASD, with males four times more likely than females to be affected.^1,2^ASD usually manifests in infants and children and presents a wide range of symptoms that vary in intensity from person to person. The heterogeneity of ASD presents a major diagnostic challenge, with clinicians typically employing a combination of lengthy parent questionnaires and clinical observation in order to evaluate children.

Standard diagnostic mechanisms for ASD are often accompanied by a range of issues that result in long waiting times for results.^3–5^ However, in recent years, significant strides have been made in the fields of computer vision and mobile technology, giving rise to the possibility of utilizing home videos of a child’s natural behaviors in order to identify characteristics linked with ASD and enable a more accurate and timely diagnosis.^6^

We previously created a mobile application called GuessWhat, which yields video data of children engaged in socially-motivated gameplay with parents in a natural home environment.^7–12^ The application presents a charades game, encouraging kids to act out a series of given prompts, such as emotions, sports, or chores. During a game, parents will open the GuessWhat application and place the smartphone on their foreheads, with the front-facing camera pointing at the child; the child then proceeds to act out the prompt displayed on the device, while the parent attempts to predict the answer (Figure 1). The game ends when the 90-second time limit is exceeded. At this point, the parent can view the video recording of the child and is then given the option to share this data with our research team.

**Fig. 1.**
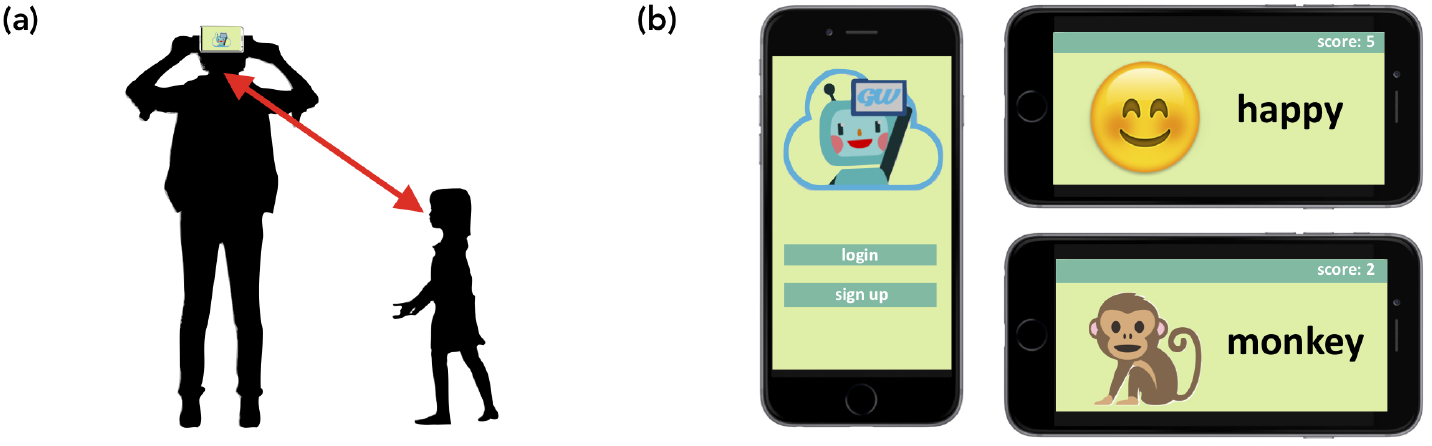
*GuessWhat Mobile Application*. (a) The parent places the mobile phone in a fixed location, allowing for the generation of a semi-structured video of gameplay. (b) The children are presented with a variety of charades prompts, such as emotions and animals.

The data collection pipeline employed by GuessWhat provides a number of benefits that make the obtained information amenable to computational analysis. First, although children are performing varied tasks in diverse environments, GuessWhat videos contain some inherent structure, with factors such as the position of the phone camera, the location of the child relative to the camera, and the game-based social interaction between the parent and child remaining generally consistent between videos. In addition, since children are in a home environment and are unencumbered by bulky hardware such as eye trackers or head mounts, they can interact with their parent and their surroundings in a natural manner. As a result, we hypothesize that computer vision algorithms can be designed to monitor socially-motivated facial engagement in children during gameplay, allowing effective identification of behaviors, eye contact events, and social interactions potentially correlated with the ASD phenotype.

In this work, we utilize computational techniques to analyze these videos and identify differences in social interaction between children with ASD and neurotypical (NT) children. We specifically focus our analysis on two common social engagement signals that are included in standard clinical diagnostic instruments and can be identified through computer vision methodologies: (1) gaze fixation patterns, which represent the regions of an individual’s visual focus, and (2) visual scanning methods, which refer to the ways in which individuals scan their surrounding environment. We perform these tasks without sharing participant videos or private patient information with human annotators.

Ultimately, the development of this system can help improve diagnosis of ASD through automated detection of impaired social interactions, mitigating the problems associated with limited diagnostic resources for neurodevelopmental disorders, especially in regions where access to care is limited.^13^ This work also demonstrates the utility of game-based approaches and automated labeling methods in preserving privacy, generating large diagnostic datasets, and improving human understanding of complex conditions.

## 2. Background

Researchers have demonstrated the utility of video data in providing diagnostic insights into gaze and engagement behaviors associated with ASD. Prior work can generally be divided into three categories: (1) manual annotation methods, (2) eye-tracking systems, or (3) use of structured environments.

### Manual Annotation Methods

Some studies have utilized human annotators to label social interaction and engagement information in video frames. Several prior works, such as those by Tariq et al. and Leblanc et al, performed manual annotation of behavioral features in home videos, which enabled the creation of classifiers that could identify ASD with high accuracy.^14–19^ Chorianopoulou et al. collected structured home videos from participants and had expert annotators label the dataset with the actions, emotions, gaze fixations, utterances, and overall level of engagement in each video; this information was then used to train a classifier to identify specific engagement features that could be correlated with ASD.^20^ Rudovic et al. trained a large and generalizable neural network to estimate engagement in children with ASD from different cultural backgrounds.^21^ Engagement labels were manually annotated by trained individuals. Although these methods enable the creation of human-vetted, accurate datasets, such approaches require large numbers of trained annotators when implemented at scale, which is both expensive and time-consuming. In addition, these techniques may compromise the privacy of participants by providing annotators with access to video footage, although some methods have been developed to address privacy concerns with crowdsourced annotations.^22,23^

### Eye-Tracking Systems

A number of studies have utilized eye trackers to identify patterns in gaze and engagement behaviors that may be indicative of ASD or other developmental conditions.^24–27^ Pusiol et al. showed that deep learning models trained on data collected from a head-mounted eyetracker and camera could be utilized to perform classification of idiopathic developmental disorder and fragile-X syndrome with high precision.^28^ Similarly, Riby et al. used eye trackers to show that individuals with ASD had atypical gaze patterns when watching movies and cartoons.^29^ In an effort to counteract artificial movements often associated with facial eye trackers, Noris et al. developed a non-intrusive eye-tracking device mounted on a hat that recorded a child’s interactions with an interviewer; the study concluded that children with ASD were more inclined to look downwards during social interaction than their NT peers.^30^ Despite the accuracy and quality of gaze data collected from such systems, eye trackers require custom hardware that can often be expensive and inaccessible, especially for individuals living in resource-limited regions. As a result, these approaches are unlikely to scale to the general population.

### Use of Structured Environments

A study by Hashemi et al. explored the use of computer vision algorithms to identify behaviors associated with ASD.^31^ A trained clinician administered a series of pre-defined, structured tasks involving toys and other visual stimuli, while a video camera captured footage of the child’s response. A computer vision system that analyzed the child’s body orientation and facial movement was able to evaluate the child’s engagement with high accuracy. Similarly, a study by Chang et al. used the front-facing camera of a mobile device to capture gaze scanning patterns as children watched strategically-designed short movies.^32^ Automated computer vision techniques were then utilized to identify differences in gaze patterns between children with ASD and neurotypical individuals. Both studies demonstrate effective methods for analyzing engagement patterns without the use of manual annotations or external eye-tracking hardware; however, both were conducted in clinical settings with highly-structured tasks and controlled environmental factors (for example, room lighting and distance of the camera from the participant’s face were fixed). As a result, the ability of these techniques to translate to natural non-clinical environments and unstructured tasks remains to be explored.

### Our Contributions

To the best of our knowledge, this is the first study that attempts to obtain diagnostic insights into ASD from mobile phone videos without the use of eye-tracking hardware, manual frame-level annotations, and structured clinical environments. We show that semi-structured home videos collected on mobile devices reveal specific regions of gaze fixation as well as visual scanning patterns that differ between individuals with ASD and neurotypical children during gameplay. With further research and development, our system can be deployed as a diagnostic tool in diverse settings at scale.

## 3. Methods

### 3.1. Data Collection

We utilized the GuessWhat mobile application to collect 449 videos of children engaged in gameplay with a parent. The participants range in age from two to fifteen and include 68 children (15 females, 53 males) who have been diagnosed with ASD as well as 27 NT children (9 females, 18 males). Each child contributed a mean of 4.7 videos (standard deviation of 7.3), resulting in a total dataset size of 1,084,267 individual frames and 11.1 hours of footage (Figure 2). All parents involved in the study consented to share their videos with our research team and completed a survey to provide the age, sex, and diagnostic status of their children.

**Fig. 2.**
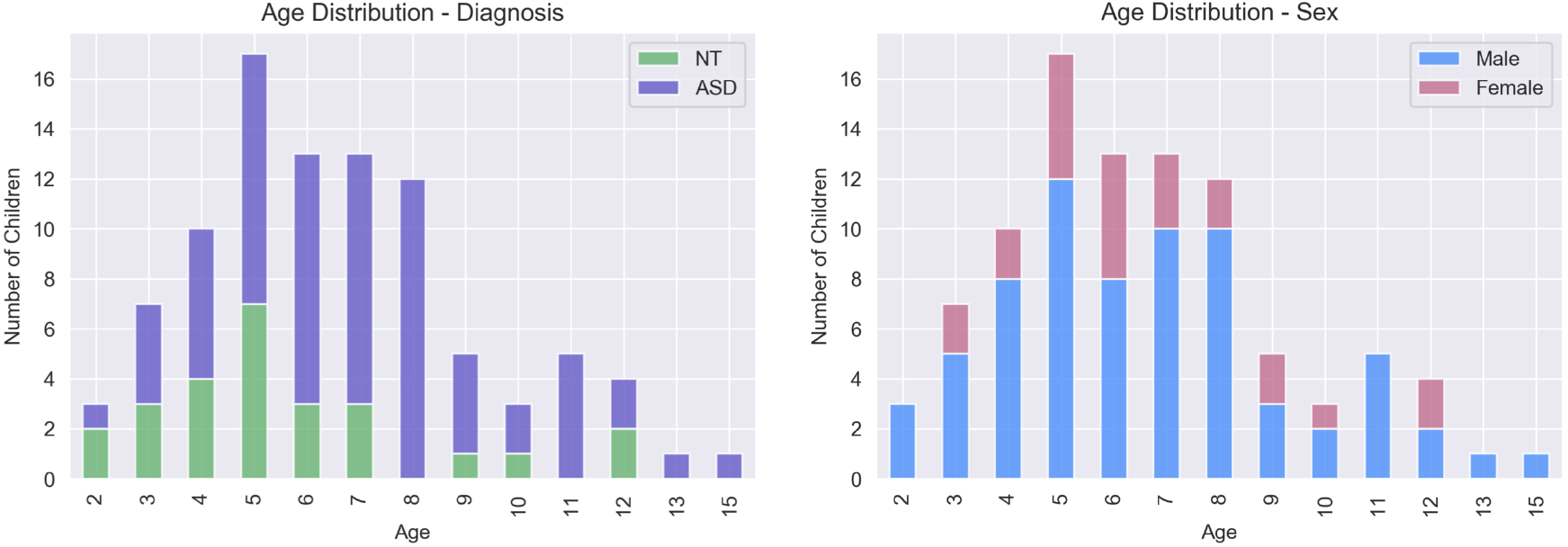
*Dataset Information*. These graphs show the breakdown of our dataset by age, diagnosis, and sex. One NT male in our dataset failed to provide his age and has been excluded from this figure.

### 3.2. Data Preprocessing

Although the semi-structured format of our video dataset presents numerous advantages, home videos are naturally heterogeneous in quality; this results in a number of challenges that must be addressed prior to computational analysis. Specifically, excessive camera movement and poor lighting conditions render some frames in our dataset too blurry for use. Also, other adults or siblings would often join in gameplay, resulting in multiple faces in the frame that make identification of the participating child challenging. Another major challenge arises from the lack of fine-grained annotations and ground truth labels; although the lack of eye-tracking hardware enables natural child motions and interactions, this also results in a lack of calibration information for obtaining accurate gaze locations.

We began our analysis with extensive quality control and data pre-processing. In order to preserve privacy, we annotated our dataset solely through the use of computational methods. We first utilized Amazon Rekognition, a powerful off-the-shelf computer vision platform developed by Amazon, to perform noisy labeling of key features in each still frame, including thirty facial landmarks and facial bounding boxes. Frames with zero or greater than two faces were removed from the dataset. We then used an open-source facial landmark annotation platform called OpenFace to obtain automated estimates of gaze directions.^33,34^ Each frame with an identifiable face was assigned a coordinate pair (*x, y*) representing the direction of the individual’s gaze. The value of *x* ranges from −1 (indicating a leftward gaze) to 1 (indicating a rightward gaze); similarly, the value of *y* ranges from −1 (indicating a downward gaze) to 1 (indicating an upward gaze) (Figure 3). Since these coordinates were assigned with respect to the smartphone camera, a frame in which an individual is gazing straight ahead into the camera is assigned a coordinate pair of (0, 0). If the OpenFace model demonstrated low confidence in gaze estimation values (defined as confidence below 75%) as a result of occluded eyes or insufficient image quality, the frame was removed from the dataset; as a result, we expect the final annotations to be of high quality, but the presence of some noise and incorrect labels is to be expected. This procedure resulted in a total of 619,620 annotated frames, representing 520,536 frames from children with ASD and 99,084 frames from NT children.

**Fig. 3.**
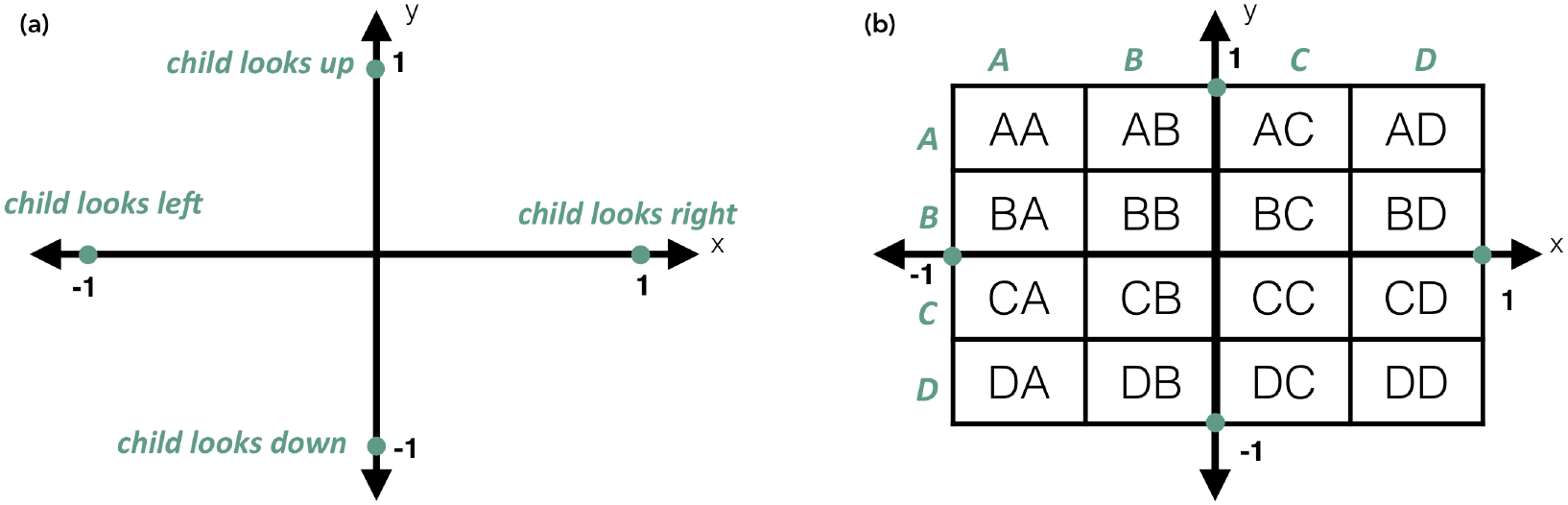
*Gaze Annotations*. (a) Gaze coordinates range between −1 and 1 on both the x-axis and y-axis. (b) In order to categorize gaze coordinates into discrete regions, we divided the gaze map into sixteen buckets. Each area of interest (AOI) is labeled with corresponding row and column letters.

Finally, in an effort to discretize gaze annotation data, we divided the coordinate map into 16 distinct areas of interest (AOIs), as shown in Figure 3. All gaze coordinates that fall within the bounds of a particular AOI are grouped together. Such an approach allows us to identify trends in an individual’s gaze fixations and scanning patterns.

### 3.3. Differential Pattern Analysis

#### 3.3.1. Gaze Fixation Patterns

Gaze fixation, which occurs when one’s gaze is held on a single target for an extended period of time, plays an important role in social interaction by signaling communicative intent and enabling interpersonal relationships. In a dyadic social interaction, individuals usually fixate their gaze on the target’s eyes. However, individuals with ASD often face difficulty with maintaining eye contact and instead tend to focus their visual attention on other regions of the target’s face. Several studies involving eye trackers and visual stimuli have shown that children with ASD tend to fixate on the mouth or other body parts; this has even been observed in children as young as two to six months of age who were later diagnosed with ASD.^35–37^ Eye contact avoidance, which is explicitly examined in standard clinical diagnostic examinations, can result in decreased facial identification and social engagement.

In order to determine the gaze fixation patterns of individuals during a single 90-second game, we utilized the coarse gaze annotations obtained from our preprocessed dataset. For each video in our dataset, we computed the percentage of time that the child fixated his or her gaze on each of the sixteen pre-defined AOIs. A two-sided permutation test was utilized at every AOI to identify statistically significant differences between the ASD and NT populations, with the null hypothesis that the fixation times for both populations followed an equivalent distribution; we calculated the difference in mean fixation times for 100,000 rearrangements of the two groups. Bonferroni correction was applied to account for multiple hypothesis tests. It is important to note that since AOIs are correlated, the Bonferroni correction is extremely stringent and will reduce the likelihood of Type 1 errors.

#### 3.3.2. Visual Scanning Patterns

Humans tend to transition their gaze between various objects in their environments when encountering visual stimuli, a phenomenon called visual scanning. The patterns and frequencies with which humans scan their surroundings can provide insight into how individuals process the world around them. In the context of social interaction, prior research has shown that individuals with ASD vary in the way that they scan a target’s facial landmarks during a social scenario, which may contribute to difficulty with interpreting emotion or nonverbal cues. This was shown by Pelphrey *et al*., who demonstrated that when presented with images of faces, NT individuals typically transitioned their gaze between core features, such as the eyes and nose, while individuals with ASD appeared to scan non-feature areas of the face, such as the forehead and cheeks.^38^ A similar study conducted by Chawarska and Shik on toddlers corroborated these findings, providing evidence of atypical scanning patterns in children with ASD when compared to their age-matched NT peers.^39^ Understanding these patterns can reveal differences in the way that individuals with ASD process visual stimuli and interact in social situations.

Modeling gaze transition patterns as a graph problem can provide insight into the regions that children focus on while scanning their environments.^40^ For each 90-second video of gameplay, we constructed a network consisting of sixteen nodes *n*_*AA*_, *n*_*AB*_, …, *n*_*DC*_, *n*_*DD*_, with each node representing a predefined AOI. When a child shifts his or her gaze between locations on the 16-AOI gaze map, an undirected edge *e* = (*n*_*i*_, *n*_*j*_) is drawn between the two corresponding nodes. Edges are weighted by the number of transitions that occur during the game. The graph can then be converted to a 16 × 16 adjacency matrix (Figure 4).

**Fig. 4.**
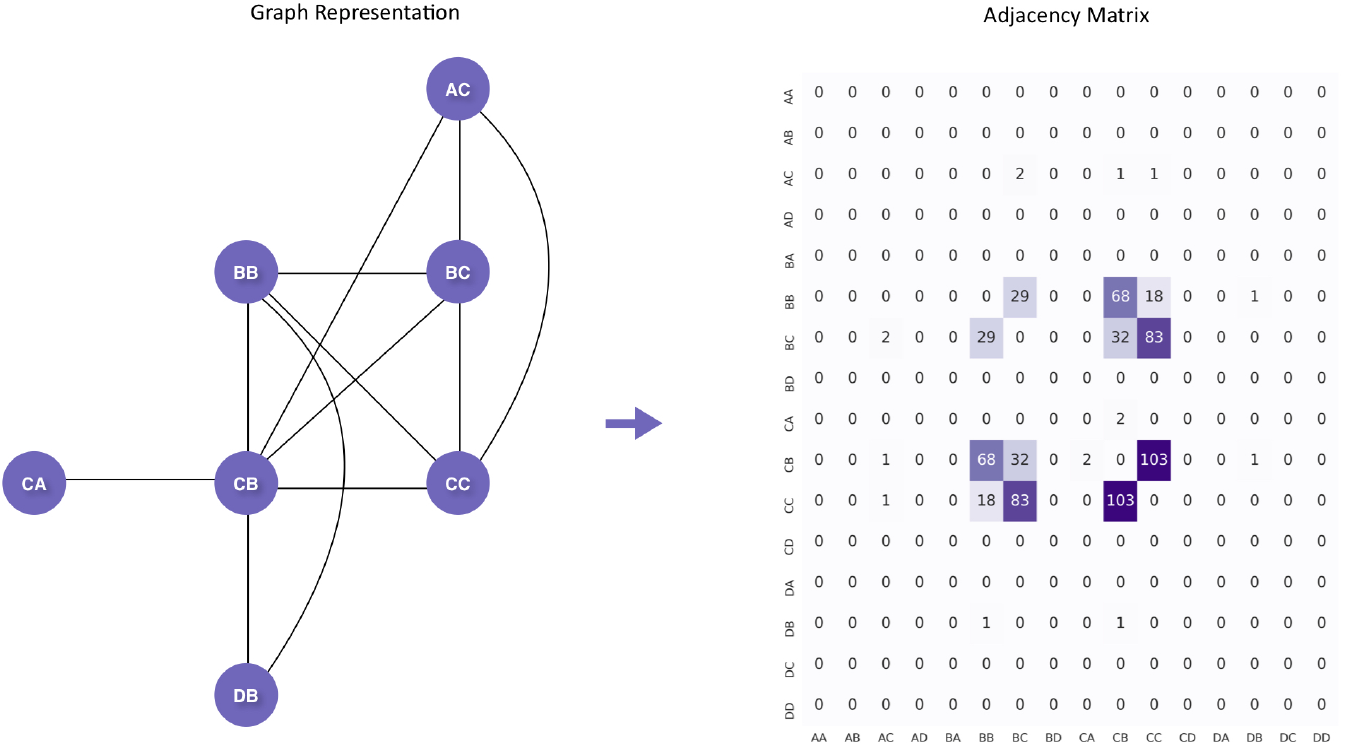
*Graph Model of Gaze Transitions*. We modeled the gaze transitions in each gameplay video as a graph, which was then used to generate a 16 × 16 adjacency matrix.

We computed adjacency matrices for all gameplay videos and normalized each matrix by dividing each entry by the total number of transitions. Then, we compute the average of all matrices associated with the NT individuals in our dataset, allowing us to obtain a single 16 × 16 matrix depicting the mean percentage of transitions occurring between each pair of AOIs in a single game. This process was repeated for the gameplay videos associated with the ASD cohort. Two-sided permutation tests were conducted at each location in the transition matrix in order to determine if there were significant differences in transition types between the two groups.

### 3.4. Deep Learning Model

Next, we utilized deep learning techniques to measure the predictive power of gaze fixation patterns. We began by formulating an approach to convert fixation data points into feature matrices that can serve as input to our classifiers. We first extracted the sequence of gaze coordinates from each video using the coarse annotation procedure described in the previous section. This resulted in a vector of *n* ordered pairs (*x,y*) for every video, where *n* represents the number of valid frames in the video and *x* and *y* are gaze fixation coordinates ranging from −1 to 1. We then matched each ordered pair with its associated AOI, as demonstrated in Figure 3. This yielded a vector of *n* AOIs, representing the regions of the gaze map that each individual fixated on during the course of a game. Next, each of the sixteen pre-defined AOIs was assigned a number from 0 to 15 in alphabetical order, with 0 representing AA and 15 representing DD; this formed a vector of *n* integers, which we will refer to as *v*.

We utilized a sliding window approach to divide *v* into separate vectors using two predefined parameters: window and shift. The window parameter *w* represents the number of frames included in a single feature vector; in our experiments, this value ranged from 50 to 500 frames, which roughly corresponds to 2 to 20 seconds of video content. The shift parameter *s* defines the number of elements by which the window slides between feature vectors, and we experimented with shift values between 10 and 100. These parameters allowed us to extract feature vectors from *v* consisting of *w* elements, with vectors separated by exactly *s* frames; note that if *s < w*, vectors will contain overlapping elements. Finally, we converted each *w*-vector into a *w* x 16 feature matrix, with each AOI integer encoded by a one-hot vector. A demonstrative example is shown in Figure 5.

**Fig. 5.**
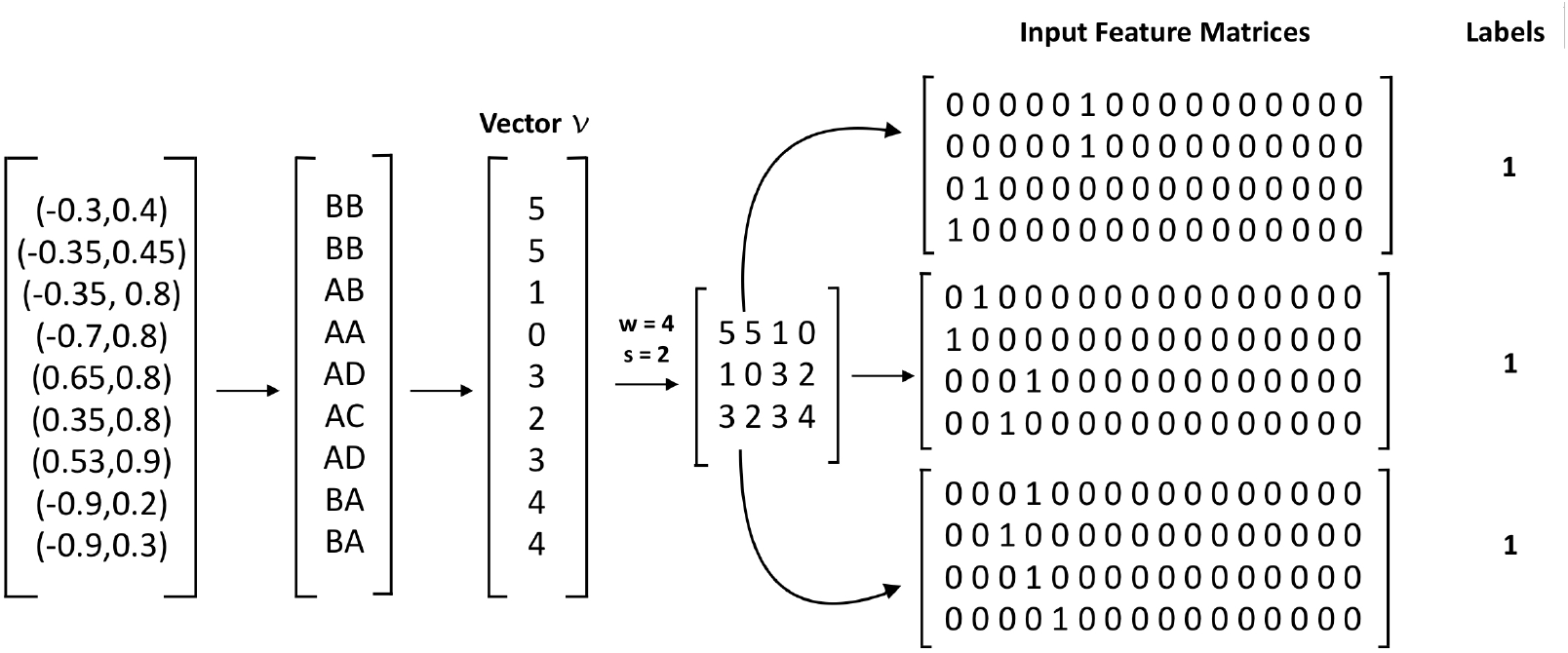
*Gaze Fixation Feature Representation*. In this demonstrative example, we begin with a video consisting of 9 frames. Gaze coordinates are matched with corresponding AOI regions. Using a window of 4 and a shift value of 2 divides vector *v* into three feature vectors. Each feature vector is then one-hot encoded. All input feature matrices are assigned the same label.

We then utilized deep learning models to determine if gaze fixation patterns could be predictive of ASD. We assigned 324 videos (275 ASD, 49 NT) in our dataset to the training set, 71 videos (62 ASD, 9 NT) to the validation set, and 54 videos (43 ASD, 11 NT) to the held-out test set, ensuring that all videos corresponding to a single child were assigned to the same set. Input feature matrices were constructed using the approach described above. A binary label *l* ∈ {0, 1} was assigned to each matrix to represent the diagnosis of the child in the associated video, with 1 representing the presence of ASD.

In order to exploit the temporal nature of our dataset, we utilized long short-term memory (LSTM) networks, which are a type of recurrent neural network that can model long-term dependencies. A *w* x 16 feature matrix served as input to an LSTM model with *w* cells; each cell accepted a one-hot encoded 16-vector as input. We used the Adam optimizer with a learning rate of 0.001, a batch size of 5, and a weighted binary cross-entropy loss function. The last cell of the LSTM network was connected to a fully-connected layer with a single-class output followed by a sigmoid non-linearity; this resulted in a final value ranging between 0 and 1. This value was rounded to the closest integer to determine the final prediction (Figure 6).

**Fig. 6.**
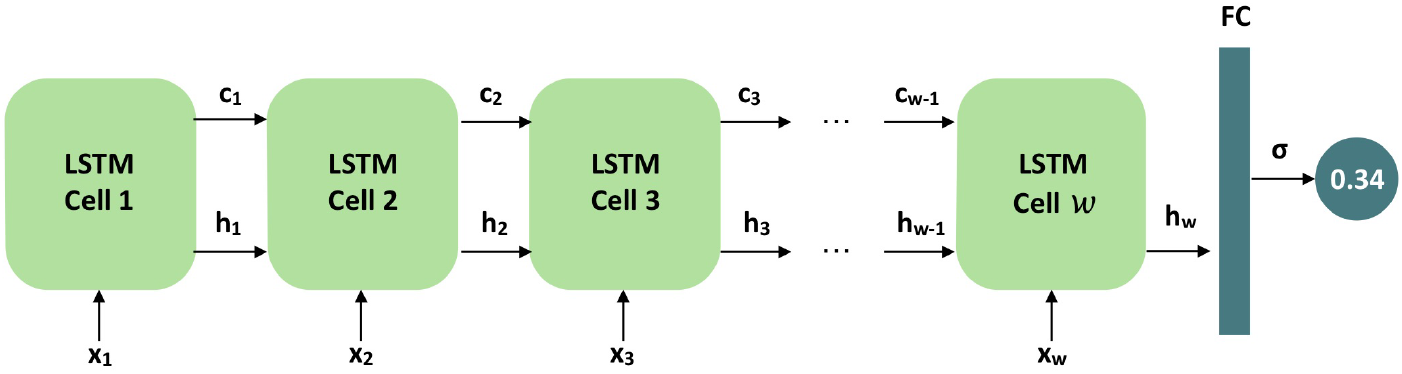
*Model Architecture*. The model consists of an LSTM network with *w* cells. Each cell accepts a one-hot vector of size 16, represented in the figure by *x*_*i*_, and outputs a cell state *c*_*i*_ and a hidden state *h*_*i*_. The final cell is connected to a fully-connected layer, which generates a single class output.

Finally, in order to characterize model performance, we report four metrics: macro-averaged (M-A) recall, macro-averaged (M-A) precision, weighted-average (W-A) recall, and weighted-average (W-A) precision. Since our dataset exhibits class imbalance with cases outnumbering controls, these metrics provide the most accurate representation of model performance. Macro-averaged statistics compute the arithmetic mean of performance on each class, while weighted-average statistics compute the weighted mean. We performed all parameter experimentation on our validation set and evaluated our final best-performing models on the held-out test set.

## 4. Results

### 4.1. Gaze Fixation Patterns Differ Between ASD and NT

We first analyzed gaze fixation patterns in order to determine if regions of focus differ between children with ASD and NT children during a single 90-second game. Coarse gaze annotations, which were obtained using the automated labeling procedure described in the Methods section, were grouped into sixteen areas of interest (AOIs), and the percentage of time that the child fixated on each region was computed. Figure 7 shows the mean percentage of time that the ASD and NT cohorts fixated on each AOI during a game. As shown by the heatmaps, children mostly fixated on the four central locations BB, BC, CB, and CC, which are located closest to the camera of the mobile phone. The distributions show that a number of differences exist between the two populations; children with ASD were most likely to fixate on locations BB and CB, while NT children spent the majority of the 90-second game focusing on locations BB and BC. We conducted a two-sided permutation test at each AOI with 100,000 permutations of the data, setting a Bonferroni-corrected significance threshold of 0.0031 in order to account for the sixteen hypothesis tests. A significant difference in fixation distributions between the two cohorts was observed at location BC (p = 0.00015) (Figure 7).

**Fig. 7.**
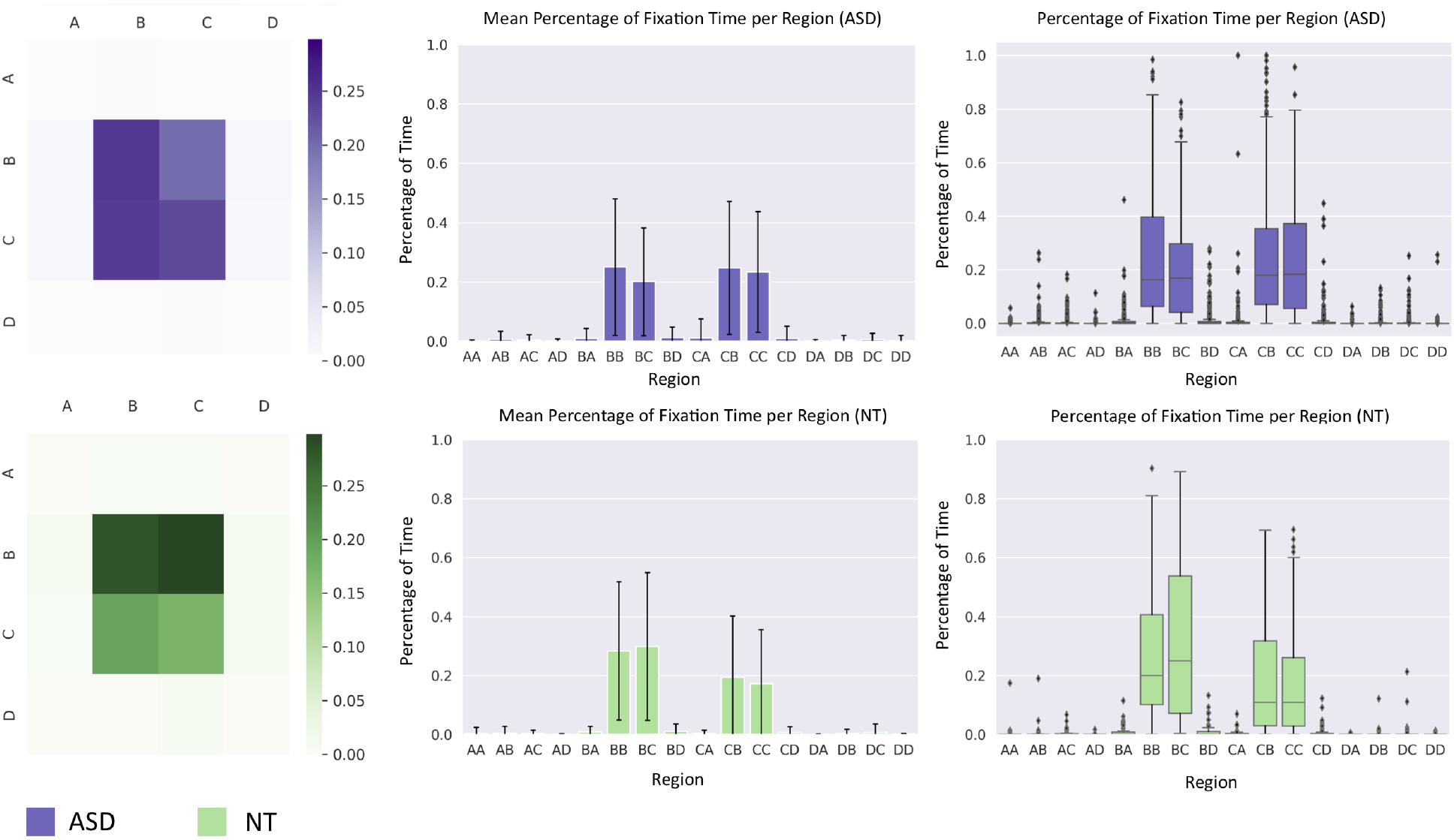
*Gaze Fixation Results*. The heatmaps located at the upper left and lower left show the mean percentage of time that an individual fixated his or her gaze on each AOI. The bar charts and box and whisker plots show the distribution of fixation times across all videos.

### 4.2. Visual Scanning Patterns Differ Between ASD and NT

Next, we utilized graph methods to analyze the ways in which participants scanned their environments during gameplay. We modeled the gaze transitions in each gameplay video as a network and computed the mean adjacency matrices for the ASD and NT populations, which are shown in Figure 8; a cell of the matrix in row *i* and column *j* represents the mean percentage of gaze transitions in a single 90-second game that occur between AOI *i* and AOI *j*. We conducted permutation tests with 100,000 permutations at each of 61 nonzero, unique locations in the adjacency matrices; since the matrix is symmetric, the distributions for each distinct transition pair were tested for significance exactly once. We then utilized a Bonferroni-corrected significance threshold of 0.0008 in order to account for 61 hypothesis tests. Our results showed a significant difference exists in the percentage of gaze transitions between regions BB and BC (p = 0.00011). As shown by the heatmaps in in Figure 8, 9.4% of the gaze transitions made by an individual with ASD occur between BB and BC; however, for NT children, 13% of the gaze transitions made during a 90 second game occur between BB and BC (Figure 8).

**Fig. 8.**
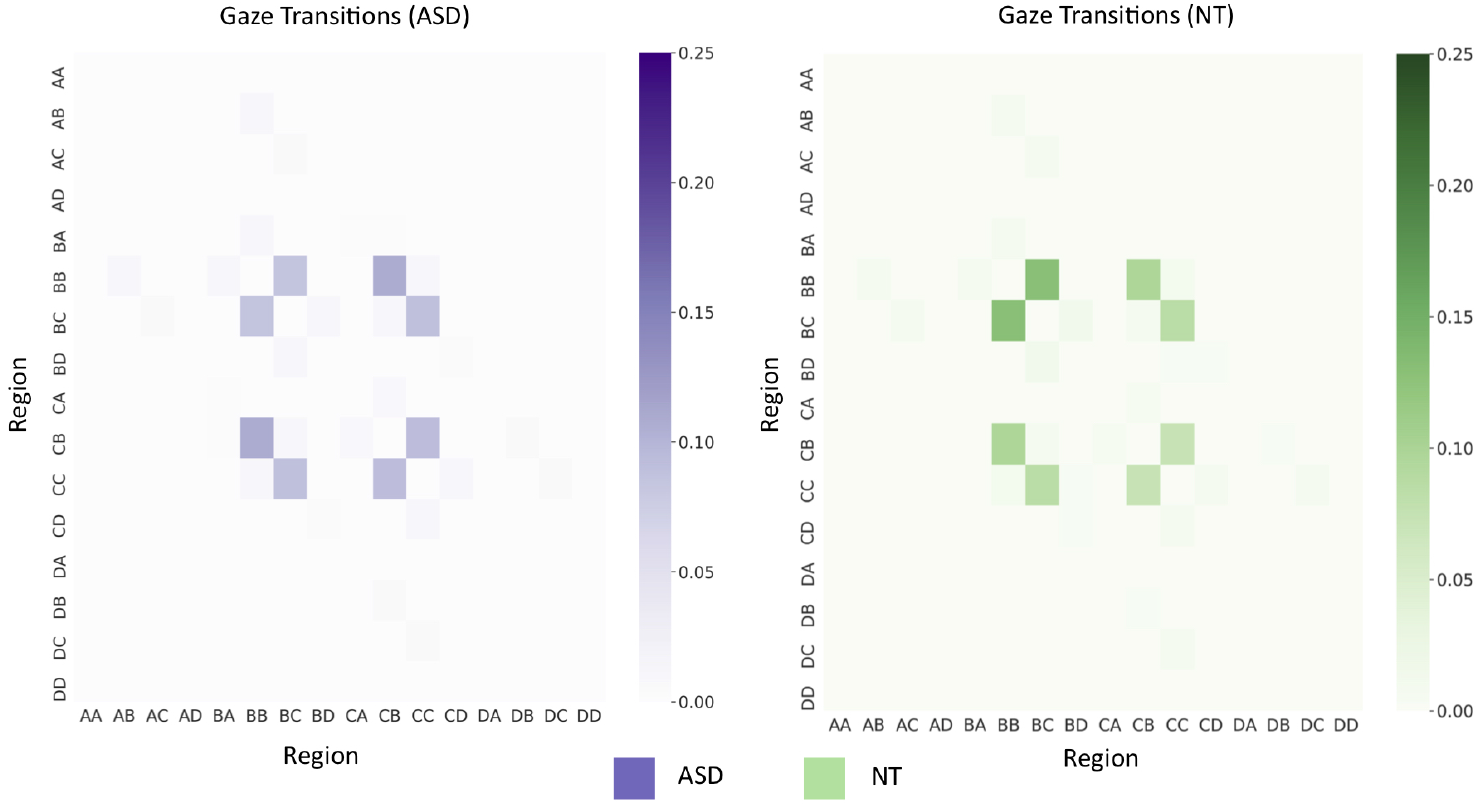
*Gaze Transition Heatmaps*. These heatmaps show the percentage of gaze transitions that occur between each pair of AOIs during a 90-second game.

### 4.3. Gaze Fixation Patterns Provide Mild Predictive Power

We measured classification performance of models trained on gaze fixation patterns. Gaze fixation coordinates were encoded as one-hot vectors and passed as input to an LSTM network, which generated a single-class output representing the likelihood of ASD. LSTM models were trained with a range of window and shift parameter values and evaluated on the validation set. Our results from the validation set allowed us to identify our top three models, which were trained with parameters (1) *w* = 100, *s* = 10, (2) *w* = 200, *s* = 10, and (3) *w* = 500, *s* = 10. These networks were then evaluated on the held-out test set; results are shown in Table 1. The model with parameters *w* = 100 and *s* = 10 demonstrated the best performance. Macro-averaged statistics are lower than weighted-average statistics, suggesting that the accuracy of prediction differs between the two classes. In summary, the results suggest that gaze fixation patterns can provide mild predictive power.

**Table 1.** *Classifier Performance on Held-Out Test Set with Gaze Fixation Features*. Precision and recall values for an LSTM model trained with the window *w* and shift *s* parameter values that achieved the best performance on the validation set.

| Window ($w$) | Shift ($s$) | M-A Recall | M-A Precision | W-A Recall | W-A Precision |
| --- | --- | --- | --- | --- | --- |
| 100 | 10 | 0.598 | 0.595 | 0.656 | 0.661 |
| 200 | 10 | 0.561 | 0.577 | 0.662 | 0.635 |
| 500 | 10 | 0.576 | 0.577 | 0.625 | 0.624 |

## 5. Discussion

In this study, we utilized computational techniques to analyze home videos and obtain diagnostic insights into ASD. We collected a large dataset of semi-structured videos featuring children engaged in gameplay with a parent, and we analyzed two key markers of social engagement that have been shown to differ between children with ASD and their neurotypical peers: (1) gaze fixation and (2) visual scanning. For each marker, we identified statistically significant differences between the two cohorts and demonstrated that this information could be useful in identifying the presence of ASD.

Our study demonstrates the potential that mobile tools hold for quantifying visual patterns and providing insights into ASD. Despite the presence of high heterogeneity and varied quality in our dataset, the automated labeling techniques and deep learning classifiers utilized in this work were able to extract usable signal and identify differences in gaze fixation and visual scanning patterns between the two cohorts. These methods also enabled us to preserve participant privacy by avoiding the use of human annotators. Our results suggest that social and visual engagement differences exist between individuals with ASD and NT individuals and that this variation can be identified through the use of mobile tools.

This work has some limitations. First, due to class imbalance in our dataset, the predictive accuracy of ASD differs from that of control individuals; this is reflected in Table 1, which shows variation between macro-averaged statistics and weighted-averaged statistics. Additional dataset augmentations will be necessary to correct this issue in the future. In addition, due to camera motion and variation in the location of the smartphone relative to the parent’s face, the gaze fixation maps are difficult to qualitatively interpret, and AOIs cannot be definitely matched to a parent’s specific facial regions.

## 6. Conclusion

Overall, this study demonstrates the utility of game-based mobile applications and heterogeneous video datasets in aiding in the diagnosis of ASD. With further research and development, the system described in this work can ultimately serve as a low-cost and accessible diagnostic tool for a global population.

## Data Availability

In order to protect participant privacy, raw videos are not made publicly available at this time.

## Acknowledgments

This work was supported by funds awarded to MV and DPW from Bio-X, the PHIND Center, and the Institute for Human-Centered Artificial Intelligence at Stanford University. MV is supported by the Knight-Hennessy Scholars program at Stanford University and the National Defense Science and Engineering Graduate (NDSEG) Fellowship. Funding sources played no role in study design; collection, analysis, and interpretation of data; writing of the manuscript; and in the decision to submit the study for publication.

## Author Contributions

MV designed the methodology and algorithms, analyzed results, and wrote the manuscript. PW, BC, and AK contributed to study design and methodology development. MV, PW, AK, and DPW verified the underlying data. PW, BC, AK, EL, KP, NS, J-YJ, MWS, and DPW provided extensive technical guidance and manuscript feedback. DPW oversaw the entire project and helped with study design and manuscript writing. All authors read and approved the final manuscript.

## References

1. E. Fombonne, The rising prevalence of autism, Journal of Child Psychology and Psychiatry 59, 717 (2018).

2. J. Baio, L. Wiggins, D. L. Christensen et al., Prevalence of autism spectrum disorder among children aged 8 years — autism and developmental disabilities monitoring network, 11 sites, united states, 2016, Surveillance Summaries 67, 1 (2018).

3. J. Bisgaier, D. Levinson, D. B. Cutts et al., Access to autism evaluation appointments with developmental-behavioral and neurodevelopmental subspecialists, Arch Pediat Adol Med 165, 673 (2011).

4. G. P. E Gordon-Lipkin, J Foster, Whittling down the wait time: Exploring models to minimize the delay from initial concern to diagnosis and treatment of autism spectrum disorder, Pediatric Clinics of North America 63, 851 (2016).

5. L. Crane, J. Chester, L. Goddard, L. Henry and E. Hill, Experiences of autism diagnosis: A survey of over 1000 parents in the united kingdom, Autism 20, 153 (2016).

6. P. Washington, N. Park, P. Srivastava, C. Voss et al., Data-driven diagnostics and the potential of mobile artificial intelligence for digital therapeutic phenotyping in computational psychiatry, Biological Psychiatry: Cognitive Neuroscience and Neuroimaging (Dec 2019).

7. H. Kalantarian, P. Washington, J. Schwartz et al., Guess what?, J. Healthc. Inform. Res..

8. H. Kalantarian, K. Jedoui, P. Washington and D. P. Wall, A mobile game for automatic emotion-labeling of images, IEEE Transactions on Games (2018).

9. H. Kalantarian, P. Washington, J. Schwartz et al., A gamified mobile system for crowdsourcing video for autism research, IEEE ICHI (2018).

10. H. Kalantarian, K. Jedoui, K. Dunlap et al., The performance of emotion classifiers for children with parent-reported autism: Quantitative feasibility study, JMIR Mental Health (2020).

11. H. Kalantarian, K. Jedoui, P. Washington et al., Labeling images with facial emotion and the potential for pediatric healthcare, Artificial intelligence in medicine (2019).

12. P. Washington, H. Kalantarian, J. Kent, A. Husic, A. Kline, E. Leblanc, C. Hou, C. Mutlu, K. Dunlap, Y. Penev, M. Varma, N. Stockham, B. Chrisman, K. Paskov, M. W. Sun, J.-Y. Jung, C. Voss, N. Haber and D. P. Wall, Training an emotion detection classifier using frames from a mobile therapeutic game for children with developmental disorders (2020).

13. M. Ning, J. Daniels, J. Schwartz, K. Dunlap, P. Washington, H. Kalantarian, M. Du and D. P. Wall, Identification and quantification of gaps in access to autism resources in the united states: An infodemiological study, J Med Internet Res 21 (Jul 2019).

14. E. Leblanc, P. Washington, M. Varma, K. Dunlap, Y. Penev, A. Kline and D. Wall, Feature replacement methods enable reliable home video analysis for machine learning detection of autism, Scientific Reports 10 (Dec 2020).

15. P. Washington, E. Leblanc, K. Dunlap, Y. Penev et al., Precision telemedicine through crowd-sourced machine learning: Testing variability of crowd workers for video-based autism feature recognition, Journal of Personalized Medicine 10 (Aug 2020).

16. Q. Tariq, S. L. Fleming, J. N. Schwartz, K. Dunlap, C. Corbin, P. Washington, H. Kalantarian, N. Z. Khan, G. L. Darmstadt and D. P. Wall, Detecting developmental delay and autism through machine learning models using home videos of bangladeshi children: Development and validation study, Journal of medical Internet research 21, p. e13822 (2019).

17. Q. Tariq, J. Daniels, J. N. Schwartz, P. Washington, H. Kalantarian and D. P. Wall, Mobile detection of autism through machine learning on home video: A development and prospective validation study, PLoS medicine 15, p. e1002705 (2018).

18. H. Abbas, F. Garberson, E. Glover and D. P. Wall, Machine learning for early detection of autism (and other conditions) using a parental questionnaire and home video screening, in 2017 IEEE International Conference on Big Data (Big Data), 2017.

19. V. A. Fusaro, J. Daniels, M. Duda, T. F. DeLuca, O. D’Angelo, J. Tamburello, J. Maniscalco and D. P. Wall, The potential of accelerating early detection of autism through content analysis of youtube videos, PLOS one 9, p. e93533 (2014).

20. Chorianopoulou, E. Tzinis, E. Iosif et al., Engagement detection for children with autism spectrum disorder, in 2017 IEEE ICASSP, 2017.

21. O. Rudovic, Y. Utsumi, J. Lee et al., Culturenet: A deep learning approach for engagement intensity estimation from face images of children with autism, in 2018 IEEE/RSJ IROS, 2018.

22. P. Washington, Q. Tariq, E. Leblanc, B. Chrisman, K. Dunlap, A. Kline, H. Kalantarian,Y. Penev, K. Paskov, C. Voss, N. Stockham, M. Varma, A. Husic, J. Kent, N. Haber, T. Winograd and D. P. Wall, Crowdsourced privacy-preserved feature tagging of short home videos for machine learning asd detection, Scientific Reports 11, p. 7620 (2021).

23. P. Washington, E. Leblanc, K. Dunlap, Y. Penev, M. Varma, J.-Y. Jung, B. Chrisman, M. W. Sun, N. Stockham, K. M. Paskov, H. Kalantarian, C. Voss, N. Haber and D. P. Wall, Selection of trustworthy crowd workers for telemedical diagnosis of pediatric autism spectrum disorder, in Biocomputing 2021,, pp. 14–25.

24. C. von Hofsten, H. Uhlig, M. Adell and O. Kochukhova, How children with autism look at events, Research in Autism Spectrum Disorders 3, 556 (2009).

25. M. Elsabbagh, E. Mercure, K. Hudry, S. Chandler, G. Pasco, T. Charman, A. Pickles, S. Baron-Cohen, P. Bolton and M. Johnson, Infant neural sensitivity to dynamic eye gaze is associated with later emerging autism, Current Biology 22, 338 (2012).

26. K. Pierce, D. Conant, R. Hazin, R. Stoner and J. Desmond, Preference for Geometric Patterns Early in Life as a Risk Factor for Autism, Archives of General Psychiatry 68, 101 (01 2011).

27. L. Yi, C. Feng, P. C. Quinn, H. Ding, J. Li, Y. Liu and K. Lee, Do individuals with and without autism spectrum disorder scan faces differently? a new multi-method look at an existing controversy, Autism Research 7, 72 (2014).

28. G. Pusiol, A. Esteva, S. S. Hall et al., Vision-based classification of developmental disorders using eye-movements, in MICCAI 2016, (Springer, 2016).

29. D. Riby and P. J. B. Hancock, Looking at movies and cartoons: Eye-tracking evidence from williams syndrome and autism., Journal of Intellectual Disability Research.

30. B. Noris, M. Barker, J. Nadel, F. Hentsch, F. Ansermet and A. Billard, Measuring gaze of children with autism spectrum disorders in naturalistic interactions, in 2011 Annual International Conference of the IEEE Engineering in Medicine and Biology Society, 2011.

31. J. Hashemi, M. Tepper, T. Vallin Spina et al., Computer vision tools for low-cost and noninvasive measurement of autism-related behaviors in infants, Autism Research and Treatment.

32. Z. Chang, J. M. Di Martino, R. Aiello, J. Baker, K. Carpenter, S. Compton, N. Davis, B. Eichner, S. Espinosa, J. Flowers, L. Franz, A. Harris, J. Howard, S. Perochon, E. M. Perrin, P. R. Krishnappa Babu, M. Spanos, C. Sullivan, B. K. Walter, S. H. Kollins, G. Dawson and G. Sapiro, Computational Methods to Measure Patterns of Gaze in Toddlers With Autism Spectrum Disorder, JAMA Pediatrics (04 2021).

33. E. Wood, T. Baltrusaitis, X. Zhang, Y. Sugano, P. Robinson and A. Bulling;, Rendering of eyes for eye-shape registration and gaze estimation, in Proceedings of the IEEE International Conference on Computer Vision (ICCV), 2015.

34. T. Baltrusaitis, A. Zadeh, Y. C. Lim and L. Morency, Openface 2.0: Facial behavior analysis toolkit, in 2018 13th IEEE International Conference on Automatic Face Gesture Recognition (FG 2018), 2018.

35. W. Jones and A. Klin, Attention to eyes is present but in decline in 2–6 month-olds later diagnosed with autism, Nature 504, 427 (12 2013).

36. K. B. Schauder, W. J. Park, Y. Tsank et al., Initial eye gaze to faces and its functional consequence on face identification abilities in autism spectrum disorder, J Neurodev Disord 11, p. 42 (2019).

37. D. Neumann, M. Spezio et al., Looking you in the mouth: abnormal gaze in autism resulting from impaired top-down modulation of visual attention, Social cognitive and affective neuroscience 1, 194 (2006).

38. K. A. Pelphrey, N. J. Sasson, J. S. Reznick et al., Visual scanning of faces in autism, J Autism Dev Disord 32, 249 (2002).

39. K. Chawarska and F. Shic, Looking but not seeing: atypical visual scanning and recognition of faces in 2 and 4-year-old children with autism spectrum disorder, Journal of Autism and Developmental Disorders 39, p. 1663 (2009).

40. Q. Guillon, M. H. Afzali, B. Rogé, S. Baduel, J. Kruck and N. Hadjikhani, The importance of networking in autism gaze analysis, PLOS ONE 10, e0141191.(10 2015).

